## Supplementary Information for "Metagenomic next-generation sequencing to characterize etiologies of non-malarial fever in a cohort living in a high malaria burden area of Uganda"

#### Table of Contents

Supplementary Results
Supplementary Tables 1-8
Supplementary Figures 1-8

#### Supplementary Results

*SARS-CoV-2 phylogenetic analysis (Figure 5)*

We performed phylogenetic inference of the 9 full genomes along with other recent SARS-CoV-2 genomes obtained from GISAID from the region and globally. The 9 full genomes represented multiple variant-of-concern and variant-of-interest lineages, including the Delta (5), Eta (3), and Alpha (1) variants. Four of the Delta-lineage viruses fell within a polytomous clade defined by a C10977T mutation. Two were identical (hCoV-19/Uganda/IDRC-CZB-04/2021 and hCoV-19/Uganda/IDRC-CZB-06/2021) and were obtained from individuals who share a household, and a third (hCoV-19/Uganda/IDRC-CZB-09/2021) had an additional 3 unique mutations. The fourth (hCoV-19/Uganda/IDRC-CZB-07/2021) clustered with two other sequences sampled from Uganda, although the most genetically similar virus within this clade had a divergence of 5 nucleotide mutations from hCoV-19/Uganda/IDRC-CZB-07/2021. The fifth Delta-lineage sequence grouped within a different clade of Delta-lineage viruses defined by a G19117T mutation. Amongst the three Eta-lineage viruses, two grouped together (hCoV-19/Uganda/IDRC-CZB-01/2021 and hCoV-19/Uganda/IDRC-CZB-02/2021), sharing C4570A, C13536T, C21811T mutations. These two viruses were separated by a C21846T mutation that was unique to hCoV-19/Uganda/IDRC-CZB-01/2021. The third Eta-lineage virus (hCoV-19/Uganda/IDRC-CZB-05/2021) was six nucleotides diverged from its nearest neighbor in the tree. The one Alpha-lineage virus, hCoV-19/Uganda/IDRC-CZB-03/2021, grouped together with other viruses sampled from Uganda (clade defined by mutations A2563G and G5992A) but was still 10 nucleotides diverged from its nearest neighbor in the tree. The estimated evolutionary rate was $7.74 \times 10^{-4}$ substitutions per site per year.

*Influenza A virus phylogenetic analysis (Figure 6)*

All 9 HA sequences generated in this study were grouped together in a clade defined by C1577T. The three most basal viruses within the clade (A/Uganda/01/2021, A/Uganda/03/2021, and A/Uganda/04/2021) were identical to each other (the latter two of which were obtained from individuals who share a household) and to a third virus sampled from Zambia. Within the clade, our sequences clustered into smaller groups as well. Two identical sequences (A/Uganda/08/2021 and A/Uganda/09/2021), obtained from individuals who share a household, were separated from the basal virus by 2 mutations (C641T and C1679T). Another pair of identical sequences (A/Uganda/05/2021 and A/Uganda/06/2021) were separated from the basal virus by 5 mutations (A453C, T206C, A353G, A797G and G862A). Finally, two terminal sequences had 1 and 3 additional unique mutations, respectively, on top of the basal genotype of this clade. The estimated evolutionary rate was $3.92 \times 10^{-3}$ substitutions per site per year.

*RSV phylogenetic analysis (Figure 7)*

The 9 RSV sequences generated as part of this study fell into 3 distinct clades. Clade 1 contained 4 of the 9 viruses. One identical pair of sequences on Clade 1 (hRSV/A/Uganda/IDRC-CZB-01/2021 and hRSV/A/Uganda/IDRC-CZB-02/2021) were obtained from individuals who share a household. Another identical pair of sequences on this clade (hRSV/A/Uganda/IDRC-CZB-07/2021 and hRSV/A/Uganda/IDRC-CZB-08/2021) was separated from the basal virus in this clade by 4 additional mutations (C2670T, T7962C, G10144A and C12109T). The inferred date for the most recent common ancestor for Clade 1 was January 2021 (95% CI: December 2020 to January 2021). Clade 1 was most closely related to samples from South Africa but still quite divergent, with 11 nucleotide mutations separating the inferred most recent common ancestor from the most basal Ugandan samples. The Ugandan and South African samples shared a common ancestor that likely circulated in January 2020 (95% CI: November 2019 to September 2020). Clade 2 contained 3 of the 9 viruses, two of which were identical (hRSV/A/Uganda/IDRC-CZB-05/2021 and hRSV/A/Uganda/IDRC-CZB-06/2021) and a third, hRSV/A/Uganda/IDRC-CZB-09/2021, with an additional C4616T mutation. The inferred date for the most recent common ancestor for Clade 2 was May 2021 (95% CI: April 2021 to June 2021). Clade 3 contained the remaining 2 viruses, hRSV/A/Uganda/IDRC-CZB-03/2021 and hRSV/A/Uganda/IDRC-CZB-04/2021, which were identical. The inferred date for the most recent common ancestor for Clade 3 was February 2021 (95% CI: January 2021 to February 2021). The estimated evolutionary rate was $8.60 \times 10^{-4}$ substitutions per site per year.

#### Supplementary Tables

##### Supplementary Table 1: Protocol details for the two processing/sequencing batches.

| | Batch 1 | Batch 2 |
|---|---|---|
| **Sample collection dates** | December 14, 2020 to February 6, 2021 | February 8, 2021 to August 22, 2021 |
| **Number of swab samples collected (how many passed QC)** | 65 (62) | 232 (232) |
| **Number of plasma samples collected (how many passed QC)** | 76 (76) | 218 (216) |
| **Number of water controls used to establish background model** | 22 | 84 |
| **Dilution of FastSelect -rRNA HMR for human RNA ribosomal depletion** | 1:100 | 1:10 |
| **Sequencer used** | Illumina NextSeq 2000 | Illumina NovaSeq 6000 |
| **Mean paired-end reads per sample** | 5.2 million | 9.2 million |

**Supplementary Table 2: Accession numbers for SARS-CoV-2, Influenza A virus, and RSV sequences.**

| **SARS-CoV-2** | |
|---|---|
| **Virus Name** | **GISAID EpiCoV Accession ID** |
| hCoV-19/Uganda/IDRC-CZB-01/2021 | EPI_ISL_8766868 |
| hCoV-19/Uganda/IDRC-CZB-02/2021 | EPI_ISL_8766869 |
| hCoV-19/Uganda/IDRC-CZB-03/2021 | EPI_ISL_8766870 |
| hCoV-19/Uganda/IDRC-CZB-04/2021 | EPI_ISL_8766871 |
| hCoV-19/Uganda/IDRC-CZB-05/2021 | EPI_ISL_8983215 |
| hCoV-19/Uganda/IDRC-CZB-06/2021 | EPI_ISL_8766872 |
| hCoV-19/Uganda/IDRC-CZB-07/2021 | EPI_ISL_8766873 |
| hCoV-19/Uganda/IDRC-CZB-08/2021 | EPI_ISL_8766874 |
| hCoV-19/Uganda/IDRC-CZB-09/2021 | EPI_ISL_8766875 |
| **Influenza A** | |
| **Virus Name** | **GISAID EpiFlu Accession ID** |
| A/Uganda/01/2021 | EPI_ISL_13493341 |
| A/Uganda/02/2021 | EPI_ISL_14016105 |
| A/Uganda/03/2021 | EPI_ISL_14016106 |
| A/Uganda/04/2021 | EPI_ISL_14016107 |
| A/Uganda/05/2021 | EPI_ISL_14016108 |
| A/Uganda/06/2021 | EPI_ISL_14016109 |
| A/Uganda/07/2021 | EPI_ISL_14016110 |
| A/Uganda/08/2021 | EPI_ISL_14016111 |
| A/Uganda/09/2021 | EPI_ISL_14016112 |
| **RSV** | |
| **Virus Name** | **GISAID EpiRSV Accession ID** |
| hRSV/A/Uganda/IDRC-CZB-01/2021 | EPI_ISL_14018007 |
| hRSV/A/Uganda/IDRC-CZB-02/2021 | EPI_ISL_14018008 |
| hRSV/A/Uganda/IDRC-CZB-03/2021 | EPI_ISL_14039044 |
| hRSV/A/Uganda/IDRC-CZB-04/2021 | EPI_ISL_14039045 |
| hRSV/A/Uganda/IDRC-CZB-05/2021 | EPI_ISL_14039046 |
| hRSV/A/Uganda/IDRC-CZB-06/2021 | EPI_ISL_14039047 |
| hRSV/A/Uganda/IDRC-CZB-07/2021 | EPI_ISL_14039048 |
| hRSV/A/Uganda/IDRC-CZB-08/2021 | EPI_ISL_14039049 |
| hRSV/A/Uganda/IDRC-CZB-09/2021 | EPI_ISL_14039050 |

**Supplementary Table 3: Classification of viral microbes/pathogens detected.**

| Viral species name | Other name(s) | Category for this analysis | Known human pathogen? |
|---|---|---|---|
| Cardiovirus B | | | -- |
| Enterovirus A | | Enterovirus | Respiratory |
| Enterovirus B | | Enterovirus | Respiratory |
| Hepatitis GB virus B | | | -- |
| Human betaherpesvirus 5 | | | -- |
| Human bocavirus | | | -- |
| Human coronavirus HKU1 | HCoV-HKU1 | Seasonal CoV | Respiratory |
| Human coronavirus NL63 | HCoV-NL63 | Seasonal CoV | Respiratory |
| Human coronavirus OC43 | HCoV-OC43 | Seasonal CoV | Respiratory |
| Human mastadenovirus C | | Adenovirus | Respiratory |
| Human metapneumovirus | | Metapneumovirus | Respiratory |
| Human orthopneumovirus | Respiratory syncytial virus (RSV) | RSV | Respiratory |
| Human orthorubulavirus 2 | Human parainfluenza virus 2 | Parainfluenza virus | Respiratory |
| Human polyomavirus 3 | | | -- |
| Human respirovirus 1 | Human parainfluenza virus 1 | Parainfluenza virus | Respiratory |
| Human respirovirus 3 | Human parainfluenza virus 3 | Parainfluenza virus | Respiratory |
| Influenza A virus | | Influenza A virus | Respiratory |
| Mamastrovirus 1 | | | -- |
| Norwalk virus | Norovirus | Norovirus | Gastrointestinal |
| Pegivirus A | | | -- |
| Pegivirus C | | | -- |
| Rhinovirus A | | Rhinovirus | Respiratory |
| Rhinovirus B | | Rhinovirus | Respiratory |
| Rhinovirus C | | Rhinovirus | Respiratory |
| Rotavirus A | | Rotavirus | Gastrointestinal |
| Severe acute respiratory syndrome-related coronavirus | SARS-CoV-2 | SARS-CoV-2 | Respiratory |

**Supplementary Table 4: Frequency table of binary results for *Plasmodium falciparum* malaria by mNGS and qPCR for the 292 plasma samples tested by mNGS.**

| N = 292 visits with plasma samples tested by mNGS | Malaria qPCR (-)<br><br>N = 224 visits | Malaria qPCR (+)<br><br>N = 67 visits | No malaria qPCR available<br><br>N = 1 visit |
|---|---|---|---|
| **Malaria mNGS (-) in plasma<br><br>N = 235 visits** | 190 visits | 45 visits | 0 visits |
| **Malaria mNGS (+) in plasma<br><br>N = 57 visits** | 34 visits | 22 visits | 1 visit |

#### Supplementary Table 5: 132 bacterial genera detected in plasma samples.

| Name of bacterial genus | Number of hits |
|---|---|
| Acinetobacter | 39 |
| Pseudomonas | 32 |
| Nocardioides | 27 |
| Escherichia | 26 |
| Sphingomonas | 25 |
| Micrococcus | 23 |
| Cutibacterium | 20 |
| Deinococcus | 17 |
| Phycicoccus | 11 |
| Arthrobacter | 9 |
| Staphylococcus | 9 |
| Delftia | 7 |
| Lactobacillus | 7 |
| Streptomyces | 7 |
| Methylobacterium | 6 |
| Prevotella | 5 |
| Rhodococcus | 5 |
| Veillonella | 5 |
| Actinomyces | 4 |
| Bacillus | 4 |
| Bifidobacterium | 4 |
| Clostridium | 4 |
| Klebsiella | 4 |
| Massilia | 4 |
| Neisseria | 4 |
| Streptococcus | 4 |
| Wolbachia | 4 |
| Xanthomonas | 4 |
| Anaerococcus | 3 |
| Comamonas | 3 |
| Herbaspirillum | 3 |
| Mycobacterium | 3 |
| Nostoc | 3 |
| Pantoea | 3 |
| Ralstonia | 3 |
| Rhizobium | 3 |
| Stenotrophomonas | 3 |
| Achromobacter | 2 |
| Acidovorax | 2 |
| Brevibacterium | 2 |
| Calothrix | 2 |
| Corynebacterium | 2 |
| Curtobacterium | 2 |
| Enterobacter | 2 |
| Enterococcus | 2 |
| Faecalibacterium | 2 |
| Finegoldia | 2 |
| Fusobacterium | 2 |
| Gemella | 2 |
| Gemmata | 2 |
| Hymenobacter | 2 |
| Janibacter | 2 |
| Kocuria | 2 |
| Leptotrichia | 2 |
| Mesorhizobium | 2 |
| Methylorubrum | 2 |
| Microlunatus | 2 |
| non-genus-specific reads in family Nocardioidaceae | 2 |
| Paenibacillus | 2 |
| Peptoniphilus | 2 |
| Propionibacterium | 2 |
| Salinicoccus | 2 |
| Sphingobium | 2 |
| Variovorax | 2 |
| Weissella | 2 |

| Name of bacterial genus | Number of hits |
|---|---|
| Actinoplanes | 1 |
| Agrobacterium | 1 |
| Alistipes | 1 |
| Alloprevotella | 1 |
| Bacteroides | 1 |
| Blastococcus | 1 |
| Blautia | 1 |
| Bradyrhizobium | 1 |
| Brevundimonas | 1 |
| Burkholderia | 1 |
| Campylobacter | 1 |
| Candidatus Methylomirabilis | 1 |
| Caulobacter | 1 |
| Cellulomonas | 1 |
| Chroococcidiopsis | 1 |
| Citrobacter | 1 |
| Collinsella | 1 |
| Cupriavidus | 1 |
| Cylindrospermum | 1 |
| Dietzia | 1 |
| Duncaniella | 1 |
| Epilithonimonas | 1 |
| Exiguobacterium | 1 |
| Filifactor | 1 |
| Gardnerella | 1 |
| Geitlerinema | 1 |
| Gordonia | 1 |
| Halomonas | 1 |
| Hassallia | 1 |
| Hydrogenophilus | 1 |
| Isoptericola | 1 |
| Knoellia | 1 |
| Lachnoclostridium | 1 |
| Lactiplantibacillus | 1 |
| Leifsonia | 1 |
| Leptolyngbya | 1 |
| Ligilactobacillus | 1 |
| Limosilactobacillus | 1 |
| Marmoricola | 1 |
| Mediterraneibacter | 1 |
| Meiothermus | 1 |
| Microbacterium | 1 |
| Microcoleus | 1 |
| Modestobacter | 1 |
| Moraxella | 1 |
| Muribaculum | 1 |
| Nocardia | 1 |
| non-genus-specific reads in family Lachnospiraceae | 1 |
| Novosphingobium | 1 |
| Ornithinimicrobium | 1 |
| Paraburkholderia | 1 |
| Paracoccus | 1 |
| Phocaeicola | 1 |
| Planktothrix | 1 |
| Porphyromonas | 1 |
| Pseudarthrobacter | 1 |
| Pseudonocardia | 1 |
| Psychrobacter | 1 |
| Rickettsia | 1 |
| Rothia | 1 |
| Scytonema | 1 |
| Serinicoccus | 1 |
| Serratia | 1 |
| Sphingobacterium | 1 |
| Sphingopyxis | 1 |
| Tetrasphaera | 1 |
| Tolypothrix | 1 |

### Supplementary Table 6: 289 bacterial genera detected in swab samples.

| Name of bacterial genus | Number of hits |
|---|---|
| Corynebacterium | 235 |
| Streptococcus | 196 |
| Acinetobacter | 185 |
| Staphylococcus | 183 |
| Pseudomonas | 176 |
| Neisseria | 155 |
| Haemophilus | 154 |
| Dolosigranulum | 152 |
| Moraxella | 147 |
| Micrococcus | 139 |
| Escherichia | 126 |
| Nocardioides | 111 |
| Deinococcus | 108 |
| Cutibacterium | 106 |
| Veillonella | 96 |
| Prevotella | 94 |
| Bacillus | 82 |
| Actinomyces | 77 |
| Pantoea | 69 |
| Arthrobacter | 68 |
| Klebsiella | 66 |
| Fusobacterium | 65 |
| Leptotrichia | 64 |
| Bacteroides | 63 |
| Streptomyces | 59 |
| Clostridium | 58 |
| Rhodococcus | 58 |
| Phycicoccus | 56 |
| Pasteurella | 54 |
| Psychrobacter | 49 |
| Sphingomonas | 49 |
| Lactobacillus | 47 |
| Alloprevotella | 46 |
| Bifidobacterium | 46 |
| Aggregatibacter | 42 |
| Anaerococcus | 42 |
| Schaalia | 38 |
| Vibrio | 38 |
| Campylobacter | 37 |
| Salmonella | 36 |
| non-genus-specific reads in family Lachnospiraceae | 35 |
| Actinobacillus | 32 |
| Gemella | 32 |
| Glaesserella | 32 |
| Mannheimia | 32 |
| Xanthomonas | 32 |
| Aeromonas | 31 |
| Kocuria | 30 |
| Rothia | 29 |
| Mycobacterium | 28 |
| Curtobacterium | 27 |
| Enterobacter | 25 |
| Avibacterium | 24 |
| Enterococcus | 24 |
| Gemmata | 24 |
| Methylobacterium | 24 |
| Serratia | 24 |
| Halomonas | 22 |
| Blautia | 21 |
| Exiguobacterium | 20 |
| Sphingobacterium | 20 |
| Microlunatus | 19 |
| Nocardia | 19 |
| Peptoniphilus | 19 |
| Porphyromonas | 19 |
| Suttonella | 19 |
| Weissella | 19 |
| Paenibacillus | 18 |
| Pseudoalteromonas | 18 |
| Shewanella | 18 |
| Enhydrobacter | 17 |
| Faecalibacterium | 17 |
| Treponema | 17 |
| Finegoldia | 16 |
| Rhizobium | 16 |
| Actinoplanes | 15 |
| Flavobacterium | 15 |
| Selenomonas | 15 |
| Capnocytophaga | 14 |
| Gordonia | 14 |
| Leuconostoc | 14 |
| Parabacteroides | 12 |
| Helcococcus | 11 |
| Lachnoclostridium | 11 |
| Lysinibacillus | 11 |
| Mycoplasma | 11 |
| Phocaeicola | 11 |
| Priestia | 11 |
| Stenotrophomonas | 11 |
| Delftia | 10 |
| Microbacterium | 10 |
| Modestobacter | 10 |
| non-genus-specific reads in family Nocardioidaceae | 10 |
| Yersinia | 10 |

| Name of bacterial genus | Number of hits |
|---|---|
| Curvibacter | 9 |
| Marinobacter | 9 |
| Micromonospora | 9 |
| Novosphingobium | 9 |
| Ornithobacterium | 9 |
| Pseudarthrobacter | 9 |
| Anaerostipes | 8 |
| Enterocloster | 8 |
| Hymenobacter | 8 |
| Lachnoanaerobaculum | 8 |
| Mediterraneibacter | 8 |
| Parvimonas | 8 |
| Propionibacterium | 8 |
| Ruminococcus | 8 |
| Tannerella | 8 |
| Azospirillum | 7 |
| Brachybacterium | 7 |
| Bradyrhizobium | 7 |
| Gardnerella | 7 |
| Granulicatella | 7 |
| Nostoc | 7 |
| Paracoccus | 7 |
| Photobacterium | 7 |
| Roseburia | 7 |
| Agrobacterium | 6 |
| Burkholderia | 6 |
| Coprococcus | 6 |
| Hungatella | 6 |
| Lachnospira | 6 |
| Limosilactobacillus | 6 |
| Megasphaera | 6 |
| Pedobacter | 6 |
| Collinsella | 5 |
| Erysipelothrix | 5 |
| Eubacterium | 5 |
| Hungateiclostridium | 5 |
| Kingella | 5 |
| Lautropia | 5 |
| Massilia | 5 |
| Mesorhizobium | 5 |
| Riemerella | 5 |
| Atopobium | 4 |
| Barnesiella | 4 |
| Butyrivibrio | 4 |
| Chryseobacterium | 4 |
| Citrobacter | 4 |
| Dietzia | 4 |
| Janibacter | 4 |
| Listeria | 4 |
| Methylorubrum | 4 |
| Mycobacteroides | 4 |
| Nocardiopsis | 4 |
| non-genus-specific reads in family Clostridiales Family XIII. Incertae Sedis | 4 |
| non-genus-specific reads in family Ruminococcaceae | 4 |
| Paraprevotella | 4 |
| Salinivibrio | 4 |
| Anaerobutyricum | 3 |
| Brevundimonas | 3 |
| Brucella | 3 |
| Cardiobacterium | 3 |
| Catonella | 3 |
| Cupriavidus | 3 |
| Erysipelatoclostridium | 3 |
| Gallibacterium | 3 |
| Lawsonella | 3 |
| Legionella | 3 |
| Marinomonas | 3 |
| Mycolicibacterium | 3 |
| Odoribacter | 3 |
| Peptostreptococcus | 3 |
| Pontibacter | 3 |
| Providencia | 3 |
| Saccharibacillus | 3 |
| Solobacterium | 3 |
| Sphingobium | 3 |
| Spirosoma | 3 |
| Streptobacillus | 3 |
| Tetrasphaera | 3 |
| Achromobacter | 2 |
| Aeromicrobium | 2 |
| Aminipila | 2 |
| Butyricimonas | 2 |
| Faecalibacillus | 2 |
| Herbaspirillum | 2 |
| Herbinix | 2 |
| Lactococcus | 2 |
| Ligilactobacillus | 2 |
| Macrococcus | 2 |
| Magnetospirillum | 2 |
| Mogibacterium | 2 |
| non-genus-specific reads in family Erysipelotrichaceae | 2 |
| non-genus-specific reads in family Moraxellaceae | 2 |
| non-genus-specific reads in family Prevotellaceae | 2 |
| Oribacterium | 2 |
| Ornithinimicrobium | 2 |
| Planktothrix | 2 |
| Ralstonia | 2 |
| Salinicoccus | 2 |
| Tuwongella | 2 |

| Name of bacterial genus | Number of hits |
|---|---|
| Abiotrophia | 1 |
| Acetobacter | 1 |
| Acidiphilium | 1 |
| Acidovorax | 1 |
| Actinomycetospora | 1 |
| Agreia | 1 |
| Alistipes | 1 |
| Alkalihalobacillus | 1 |
| Alteromonas | 1 |
| Alysiella | 1 |
| Amycolatopsis | 1 |
| Anaerobacillus | 1 |
| Anoxybacillus | 1 |
| Aquabacterium | 1 |
| Arcanobacterium | 1 |
| Arsenicicoccus | 1 |
| Aurantimonas | 1 |
| Aureimonas | 1 |
| Bordetella | 1 |
| Brevibacillus | 1 |
| Brevibacterium | 1 |
| Brochothrix | 1 |
| Candidatus Nitrosacidococcus | 1 |
| Carnobacterium | 1 |
| Cellulomonas | 1 |
| Cellvibrio | 1 |
| Chromobacterium | 1 |
| Chroococcidiopsis | 1 |
| Clostridioides | 1 |
| Comamonas | 1 |
| Devosia | 1 |
| Dialister | 1 |
| Dichelobacter | 1 |
| Dyadobacter | 1 |
| Dysgonomonas | 1 |
| Eikenella | 1 |
| Elizabethkingia | 1 |
| Empedobacter | 1 |
| Erwinia | 1 |
| Ewingella | 1 |
| Filimonas | 1 |
| Flammeovirga | 1 |
| Flavonifractor | 1 |
| Frigoribacterium | 1 |
| Gluconobacter | 1 |
| Glutamicibacter | 1 |
| Isoptericola | 1 |
| Janthinobacterium | 1 |
| Jeotgalicoccus | 1 |
| Johnsonella | 1 |
| Knoellia | 1 |
| Komagataeibacter | 1 |
| Kribbella | 1 |
| Kutzneria | 1 |
| Lactiplantibacillus | 1 |
| Leclercia | 1 |
| Leptospira | 1 |
| Luteimonas | 1 |
| Lysobacter | 1 |
| Mammaliicoccus | 1 |
| Megamonas | 1 |
| Methylomonas | 1 |
| Mobilicoccus | 1 |
| Mobiluncus | 1 |
| Morganella | 1 |
| Muribaculum | 1 |
| Negativicoccus | 1 |
| non-genus-specific reads in family Muribaculaceae | 1 |
| non-genus-specific reads in family Pasteurellaceae | 1 |
| non-genus-specific reads in family Rhodospirillaceae | 1 |
| Oceanisphaera | 1 |
| Oligella | 1 |
| Pelosinus | 1 |
| Peptoanaerobacter | 1 |
| Phascolarctobacterium | 1 |
| Phyllobacterium | 1 |
| Planococcus | 1 |
| Pseudescherichia | 1 |
| Pseudobutyrivibrio | 1 |
| Pseudonocardia | 1 |
| Pseudoxanthomonas | 1 |
| Pusillimonas | 1 |
| Rahnella | 1 |
| Rickettsia | 1 |
| Rodentibacter | 1 |
| Rubrobacter | 1 |
| Shigella | 1 |
| Simonsiella | 1 |
| Stomatobaculum | 1 |
| Sutterella | 1 |
| Tatumella | 1 |
| Taylorella | 1 |
| Thiomonas | 1 |
| Ureaplasma | 1 |
| Variovorax | 1 |
| Virgibacillus | 1 |

**Supplementary Table 7: Household co-detection of viral pathogens, and time between infections in a household.** Days between infections in the 11 households that had multiple individuals with the same respiratory viral pathogen detected during the sampling period. HH: household.

| Household | Virus | Days since first infection in HH |
|---|---|---|
| HH_1 | Influenza A virus | 0 |
| HH_1 | Influenza A virus | 11 |
| HH_2 | Influenza A virus | 0 |
| HH_2 | Influenza A virus | 0 |
| HH_3 | Influenza A virus | 0 |
| HH_3 | Influenza A virus | 39 |
| HH_4 | Parainfluenza virus | 0 |
| HH_4 | Parainfluenza virus | 21 |
| HH_5 | Parainfluenza virus | 0 |
| HH_5 | Parainfluenza virus | 10 |
| HH_6 | Rhinovirus | 0 |
| HH_6 | Rhinovirus | 0 |
| HH_6 | Rhinovirus | 0 |
| HH_7 | Rhinovirus | 0 |
| HH_7 | Rhinovirus | 0 |
| HH_8 | Rhinovirus | 0 |
| HH_8 | Rhinovirus | 84 |
| HH_9 | RSV | 0 |
| HH_9 | RSV | 0 |
| HH_10 | SARS-CoV-2 | 0 |
| HH_10 | SARS-CoV-2 | 11 |
| HH_11 | SARS-CoV-2 | 0 |
| HH_11 | SARS-CoV-2 | 6 |

**Supplementary Table 8: Co-detections of malaria in plasma (by qPCR and/or by mNGS) and selected viral pathogenic microbes detected in plasma/NP swab (by mNGS) at a given visit.** These are for the 273 visits that had paired mNGS sample collections, as summarized in Table 3. The included viruses are those that are designated as “respiratory” or “gastrointestinal” pathogens in Supplementary Table 3.

| qPCR result (malaria) | mNGS result (malaria; plasma) | Virus(es) identified by mNGS (plasma or swab) | Number of visits |
|---|---|---|---|
| Negative | Negative | Enterovirus A | 2 |
| Negative | Negative | Enterovirus B | 2 |
| Negative | Negative | Human coronavirus NL63 | 1 |
| Negative | Negative | Human coronavirus OC43 | 2 |
| Negative | Negative | Human mastadenovirus C | 2 |
| Negative | Negative | Human metapneumovirus | 2 |
| Negative | Negative | Human orthopneumovirus | 10 |
| Negative | Negative | Human orthorubulavirus 2 | 1 |
| Negative | Negative | Human respirovirus 1 | 7 |
| Negative | Negative | Human respirovirus 3 | 2 |
| Negative | Negative | Human respirovirus 3 + Rhinovirus C | 1 |
| Negative | Negative | Influenza A virus | 8 |
| Negative | Negative | Norwalk virus | 1 |
| Negative | Negative | Rhinovirus A | 4 |
| Negative | Negative | Rhinovirus A + Rhinovirus B | 1 |
| Negative | Negative | Rhinovirus B | 4 |
| Negative | Negative | Rhinovirus C | 14 |
| Negative | Negative | Rotavirus A | 1 |
| Negative | Negative | SARS-CoV-2 | 6 |
| Negative | Positive | Human coronavirus OC43 | 1 |
| Negative | Positive | Human metapneumovirus | 4 |
| Negative | Positive | Human orthopneumovirus | 1 |
| Negative | Positive | Human respirovirus 1 | 1 |
| Negative | Positive | Rhinovirus C | 4 |
| Negative | Positive | Rotavirus A | 2 |
| Negative | Positive | Rotavirus A + Rhinovirus C | 1 |
| Negative | Positive | SARS-CoV-2 | 1 |
| Positive | Negative | Human coronavirus HKU1 | 1 |
| Positive | Negative | Human coronavirus OC43 | 1 |
| Positive | Negative | Human mastadenovirus C + SARS-CoV-2 + Human respirovirus 1 | 1 |
| Positive | Negative | Human metapneumovirus | 1 |
| Positive | Negative | Human respirovirus 1 | 1 |
| Positive | Negative | Human respirovirus 3 | 2 |
| Positive | Negative | Influenza A virus | 2 |
| Positive | Negative | Rhinovirus A | 1 |
| Positive | Negative | Rhinovirus C | 1 |
| Positive | Negative | SARS-CoV-2 | 1 |
| Positive | Positive | Enterovirus A | 2 |
| Positive | Positive | Human respirovirus 1 | 1 |
| Positive | Positive | Human respirovirus 3 | 1 |
| Positive | Positive | Influenza A virus | 1 |
| Positive | Positive | Rhinovirus B | 1 |
| Positive | Positive | Rhinovirus C | 2 |

**Supplementary Figures**

**Supplementary Figure 1: Distribution of sample input RNA mass and total reads by sample type. (A)** Input RNA in picograms. Blue line indicates 25 pg (i.e., amount of spike-in control in each sample). **(B)** Total reads per sample. These figures include the 292 plasma samples and 294 swab samples which passed CZ ID’s QC filters and were thus included in this analysis. Sample input is calculated as: (25 pg / ERCC reads) * (total reads - ERCC reads).

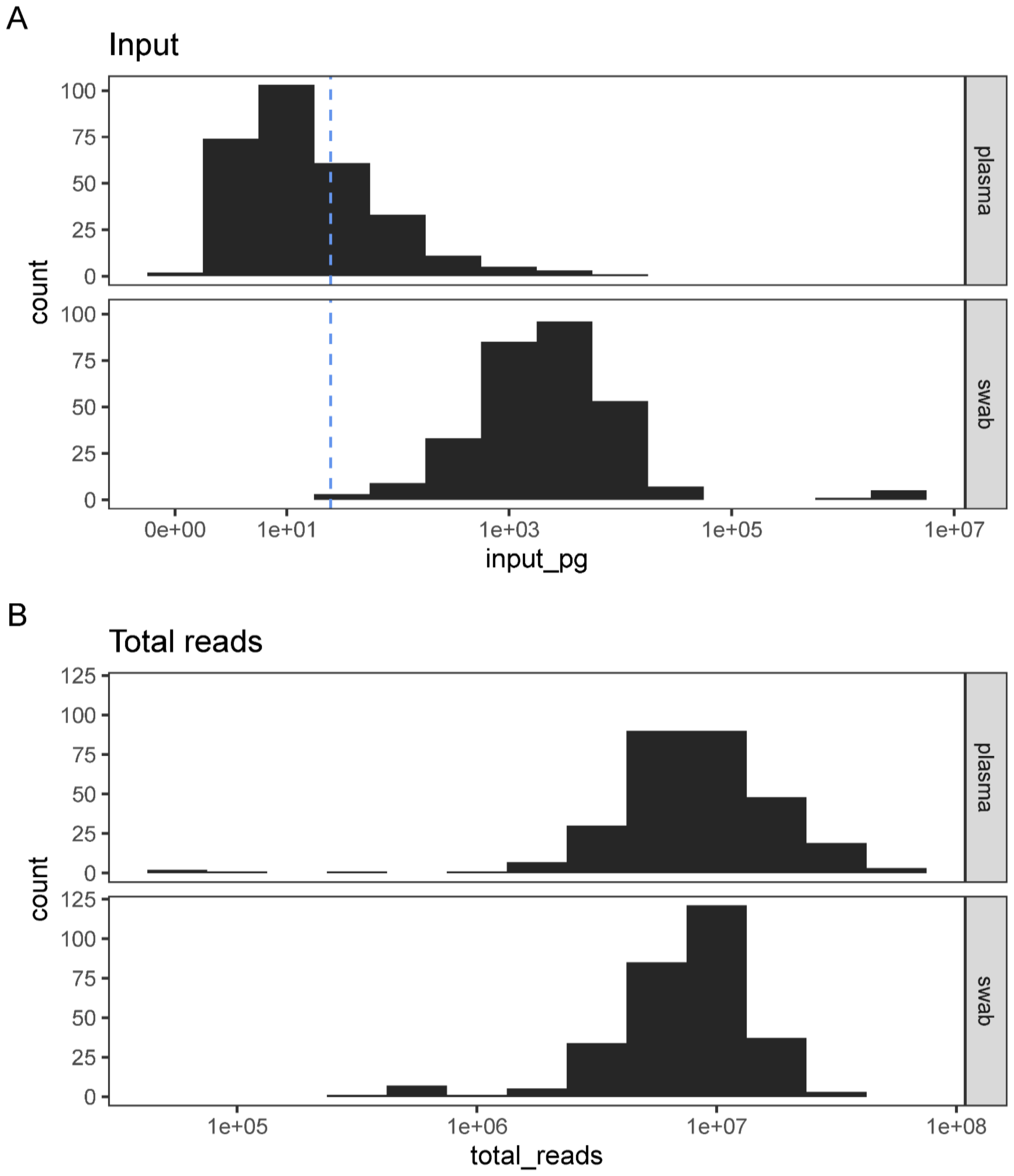

**Supplementary Figure 2: Flowchart of PRISM Border Cohort study visits and corresponding mNGS sample collection conducted in this pilot.**

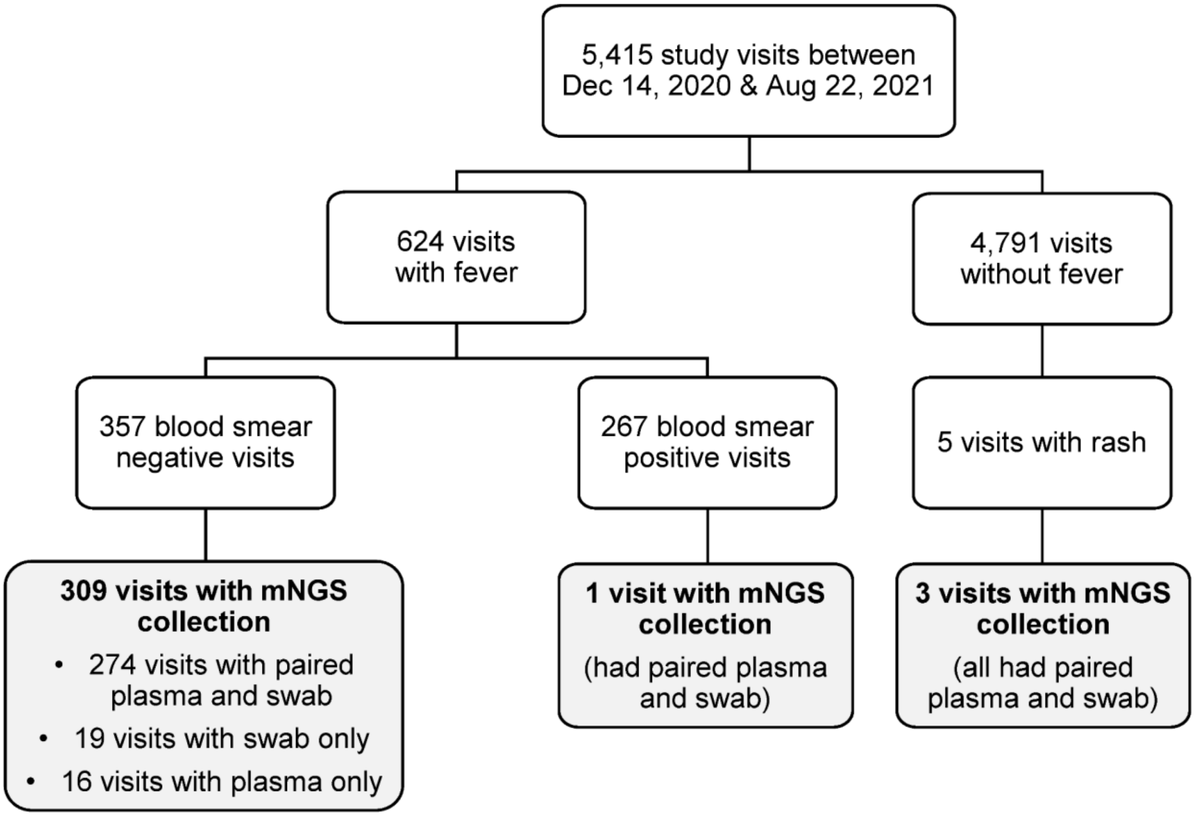

**Supplementary Figure 3: Comparison of *Plasmodium falciparum* malaria qPCR parasite densities vs. mNGS NT rPMs in plasma samples.** The filled points (which have non-zero values for both qPCR and mNGS NT rPM to *Plasmodium falciparum*) were included in a linear regression of log parasite density (y-axis) versus log NT rPM (x-axis). Note that the model includes samples that did not meet our threshold criteria to be called as positive for *Plasmodium falciparum* by mNGS (indicated by color). The open points (which have a zero value by either or both assays, and have pseudocounts added for visualization purposes) were excluded from the regression. The horizontal line indicates the threshold to be called positive for *Plasmodium falciparum* by qPCR. Estimated slope = 1.01 and adjusted $R^2$ = 0.49.

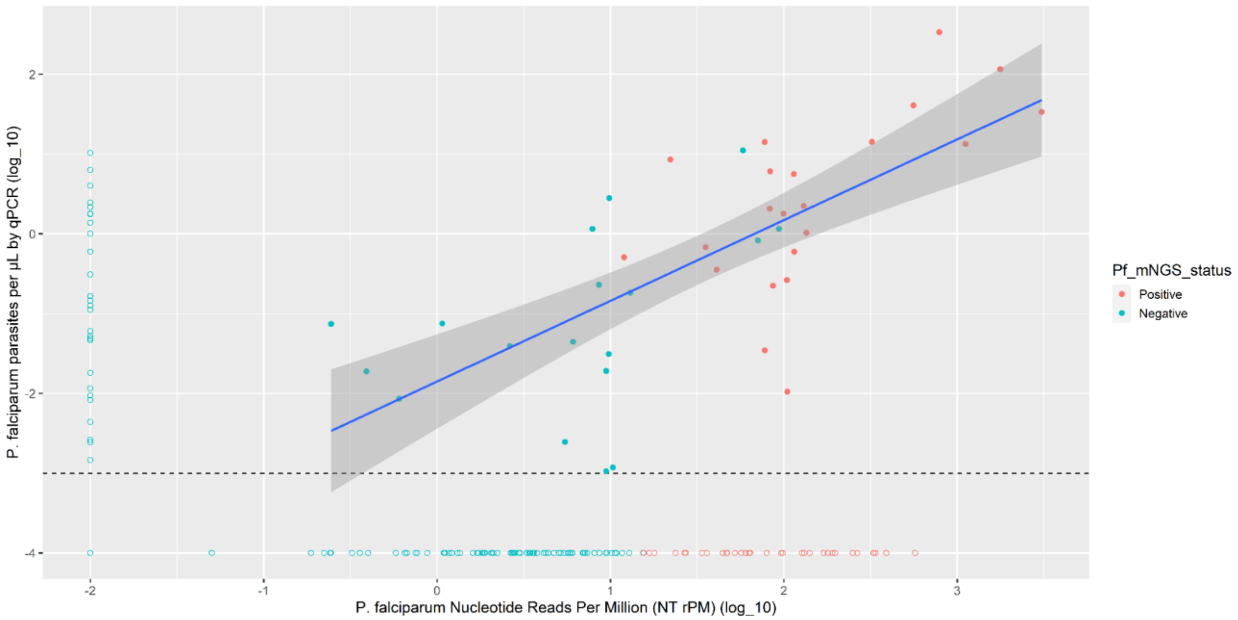

**Supplementary Figure 4: Composition of microbial detections in plasma samples.** Each column is a distinct sample. The top row shows the sample input. The 25 pg threshold is shown by the dotted red line. The y-axes of this row vary by panel. The middle row represents the proportion of reads within the sample that corresponded to each kingdom prior to filtering. The bottom row represents the proportion of reads within the sample after filtering. Only filtered reads that map to bacterial, viral, or eukaryotic species are included in filtered barplots. Panel **(A)** includes samples in the bottom half of sample inputs, and panel **(B)** includes samples in the top half of sample inputs.

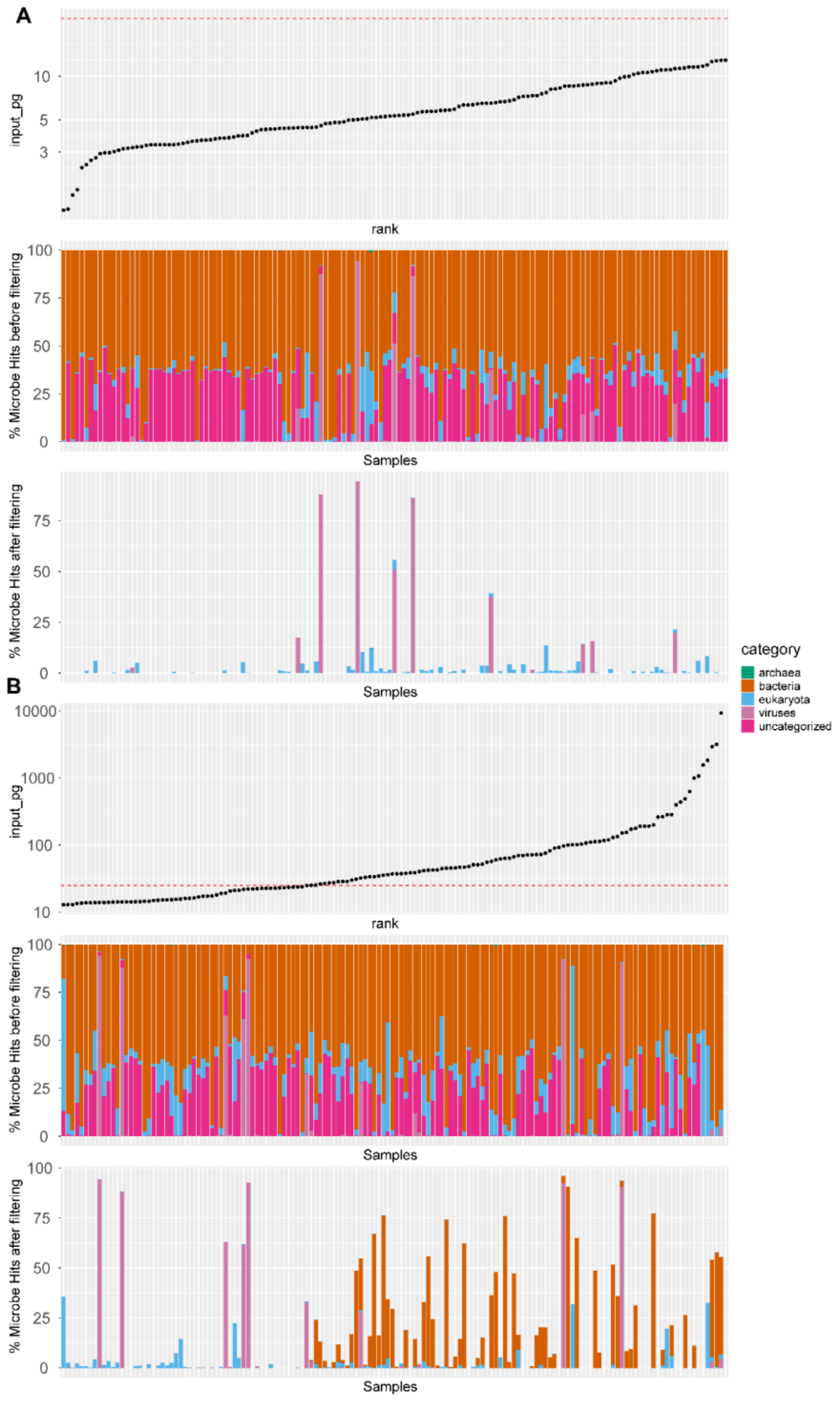

**Supplementary Figure 5: Composition of microbial detections in NP swab samples.** Each column is a distinct sample. The top row shows the sample input. The 25 pg threshold is shown by the dotted red line. The 2 samples in green denote failed ERCC, and thus not the accurate sample input. The y-axes of this row vary by panel. The middle row represents the proportion of reads within the sample that corresponded to each kingdom prior to filtering. The bottom row represents the proportion of reads within the sample after filtering. Only filtered reads that map to bacterial, viral, or eukaryotic species are included in filtered barplots. Panel **(A)** includes samples in the bottom half of sample inputs, and panel **(B)** includes samples in the top half of sample inputs.

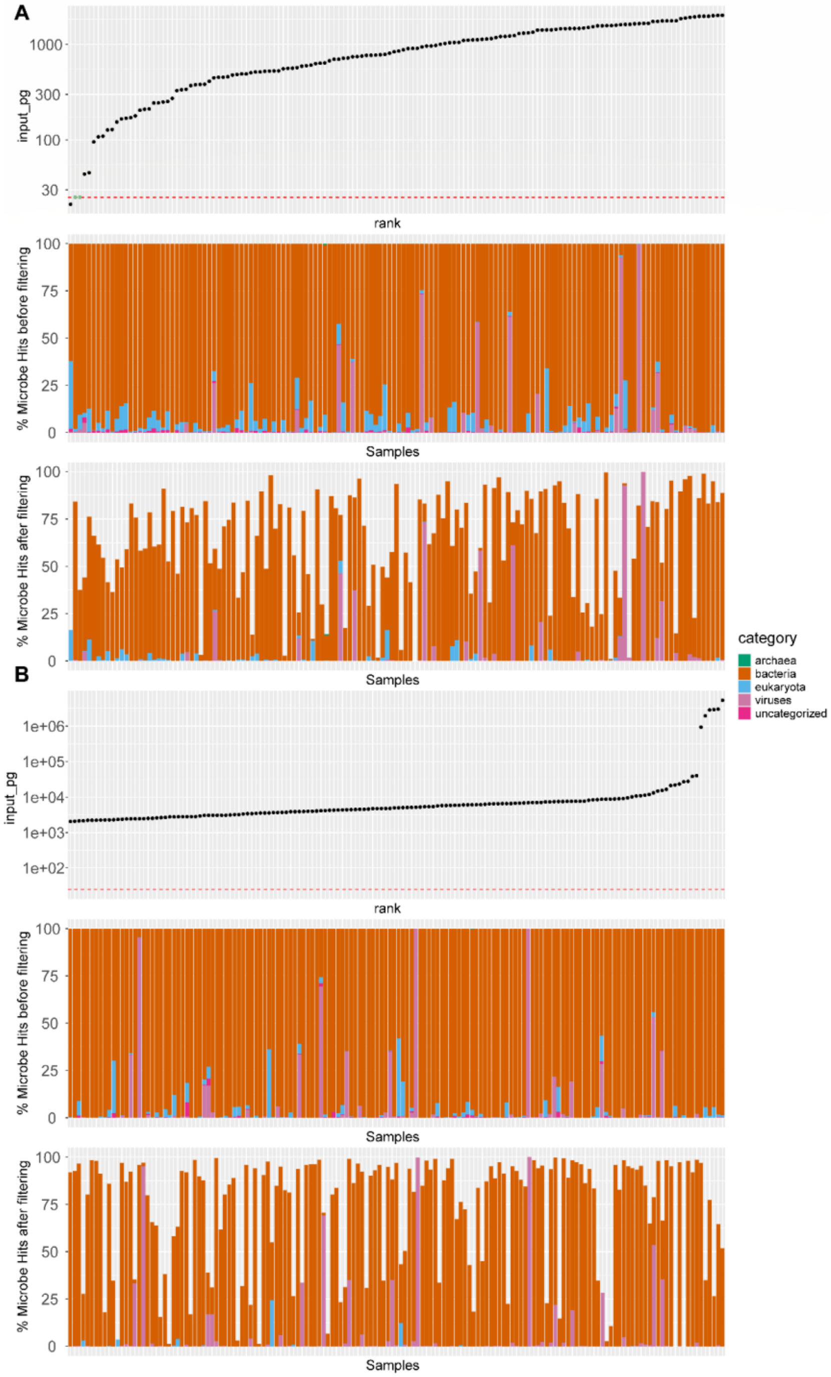

**Supplementary Figure 6: Comparison of viral microbes detected in (paired) plasma and NP swab samples.** All viral microbe species from Figure 2E are included; respiratory and gastrointestinal viral microbes that are known to be pathogenic to humans (see Supplementary Table 3) are highlighted with an asterisk.

**Viral detections**

| In 19 pairs with ≥ 1 viral detection in plasma sample only (no viral detection in swab) | In 81 pairs with ≥ 1 viral detection in swab sample only (no viral detection in plasma) |
|---|---|
| Pegivirus C (13 detections)<br>Pegivirus A (10)<br>Enterovirus A* (2)<br>Human mastadenovirus C* (2)<br>Enterovirus B* (1)<br>Hepatitis GB virus B (1)<br>Mamastrovirus 1 (1)<br>Rotavirus A* (1) | Rhinovirus C* (17 detections)<br>Human orthopneumovirus* (10)<br>Human respirovirus 1* (10)<br>Influenza A virus* (10)<br>Human metapneumovirus* (7)<br>Rhinovirus B* (6)<br>SARS-CoV-2* (6)<br>Rhinovirus A* (5)<br>Human coronavirus OC43* (4)<br>Human respirovirus 3* (4)<br>Human bocavirus (1)<br>Human coronavirus HKU1* (1)<br>Human coronavirus NL63* (1)<br>Human orthorubulavirus 2* (1) |

| In 19 unpaired plasma samples (no swab collected) | In 21 unpaired swab samples (no plasma collected) |
|---|---|
| Enterovirus A* (1 detection)<br>Human orthopneumovirus* (1)<br>Pegivirus C (1)<br>Rhinovirus C* (1) | Influenza A virus* (3 detections)<br>Human mastadenovirus C* (2)<br>Human metapneumovirus* (2)<br>Rhinovirus A* (2)<br>SARS-CoV-2* (2)<br>Human respirovirus 1* (1)<br>Rhinovirus B* (1)<br>Rhinovirus C* (1) |

**In 8 pairs with identical viral detection in plasma & swab samples**

| Pair | Detection in plasma & swab |
|---|---|
| 1 | Rhinovirus C* |
| 2 | Rhinovirus C* |
| 3 | Enterovirus B* |
| 4 | Human respirovirus 3* |
| 5 | Enterovirus A* |
| 6 | Rotavirus A* |
| 7 | Rotavirus A* |
| 8 | Rhinovirus C* |

**153 pairs with no viral detection in plasma or swab sample**

**In 12 pairs with discordant viral detection(s) in plasma & swab samples**

| Pair | Detection(s) in plasma | Detection(s) in swab |
|---|---|---|
| 1 | Enterovirus A*<br>Human betaherpesvirus 5 | Enterovirus A* |
| 2 | Human betaherpesvirus 5 | Rhinovirus C* |
| 3 | Rotavirus A* | Rhinovirus C*<br>Rotavirus A* |
| 4 | Human mastadenovirus C* | Human respirovirus 1*<br>SARS-CoV-2* |
| 5 | Pegivirus A | Rhinovirus C* |
| 6 | Pegivirus A<br>Pegivirus C | SARS-CoV-2* |
| 7 | Pegivirus C | Rhinovirus A* |
| 8 | Pegivirus C | Human respirovirus 3* |
| 9 | Pegivirus A<br>Pegivirus C | Influenza A virus* |
| 10 | Pegivirus A<br>Pegivirus C | SARS-CoV-2* |
| 11 | Cardiovirus B | Human orthopneumovirus* |
| 12 | Norwalk virus* | Human polyomavirus 3 |

**Supplementary Figure 7: Respiratory pathogen-specific prevalence among the sample population by age group.** Prevalence was estimated as the probability of detection by mNGS. We assumed that each pathogen detection was independent due to the infrequency of co-detections of respiratory viruses in this study (3 of 119 visits). The point depicts the posterior median probability and the outer interval is the 95% credible interval, using a binomial model.

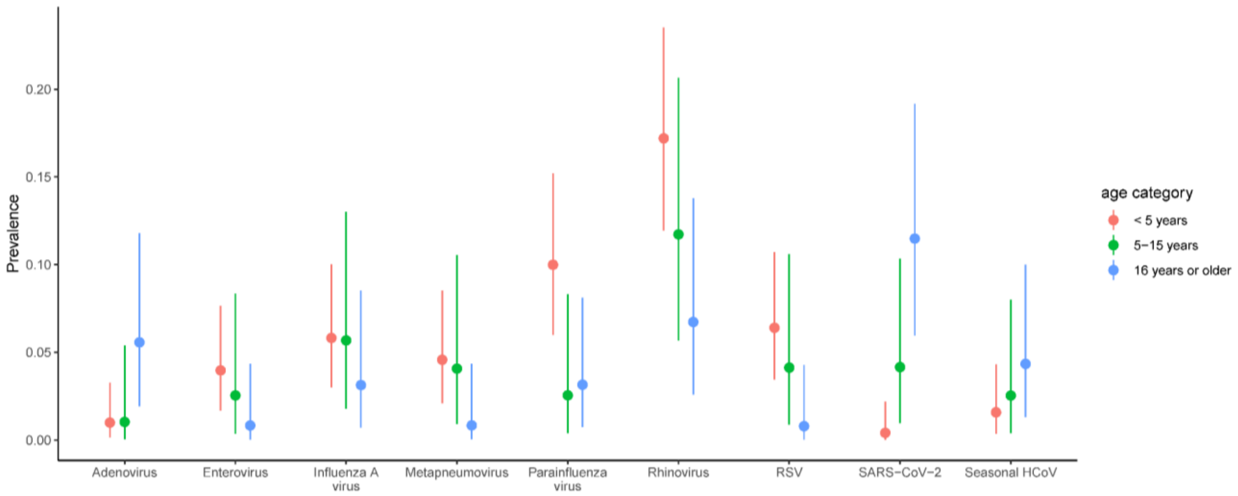

**Supplementary Figure 8: Analysis of SARS-CoV-2, Influenza A, and RSV sequences generated in this mNGS study, with respect to nucleotide divergence. (A)** 9 SARS-CoV-2 genomes from this study, with differences in the number of mutations on the x-axis. **(B)** 9 Influenza A (H3N2) HA gene segments from this study, with divergence (number of mutations per site) on the y-axis. **(C)** 9 RSV genomes from this study, with divergence (number of mutations per site) on the y-axis. Samples from the same household are shown in the same color (the samples labeled in black are from a household with a singleton sample).

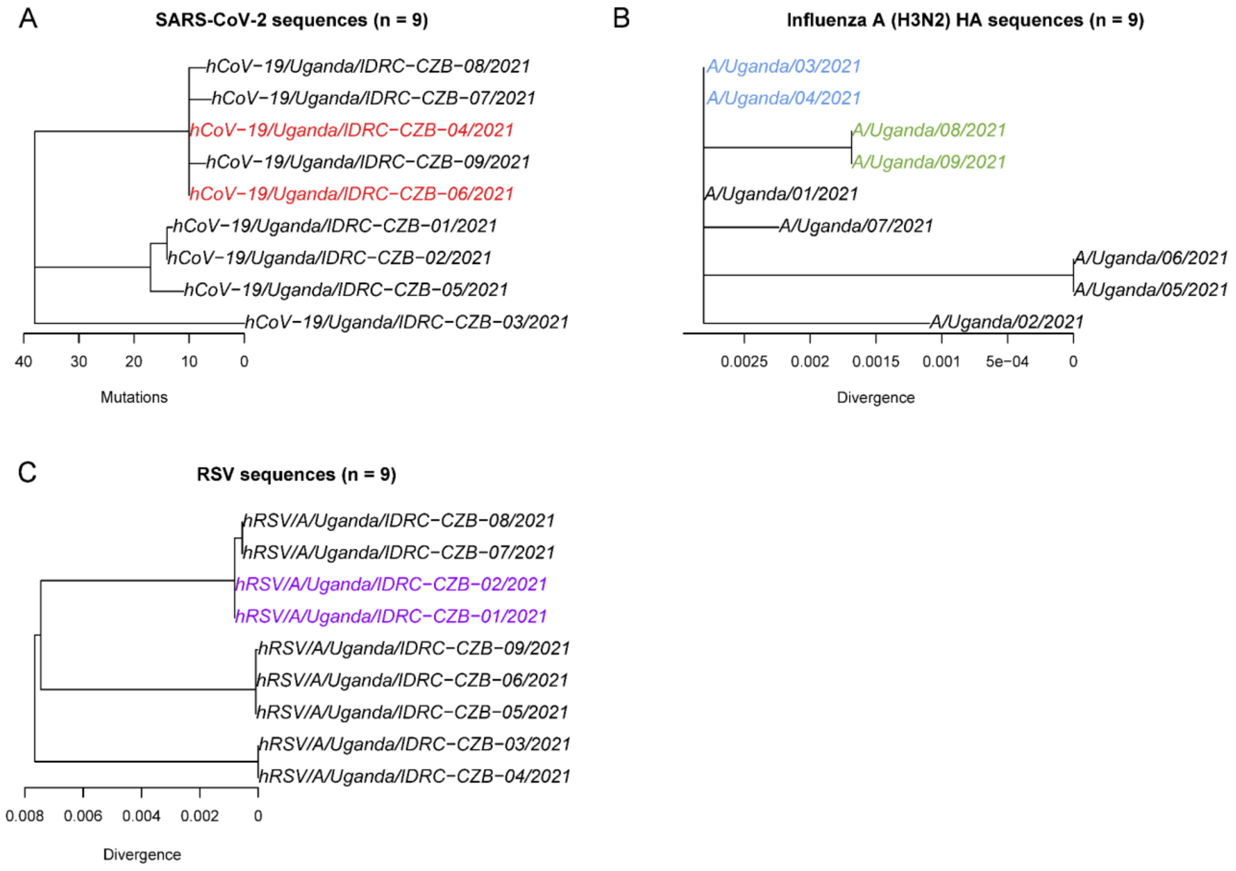
